# When the Clock Shifts: Menopause Timing is Associated with Reduced Cognitive Performance and Gray Matter Volume in a Population-Based Cohort

**DOI:** 10.1101/2025.02.22.25322720

**Authors:** Nitsan Schwarz, Daniel Harlev, Eyal Bergmann, Noham Wolpe

**Affiliations:** Department of Physical Therapy, The Stanley Steyer School of Health Professions, Faculty of Medical & Health Sciences, Tel Aviv University, Tel Aviv 6997801, Israel; Sagol School of Neuroscience, Tel Aviv University, Tel Aviv 6997801, Israel; Department of Psychiatry, Rambam Health Care Campus, Haifa, Israel; MRC Cognition & Brain sciences Unit, Department of Psychiatry, University of Cambridge, Cambridge CB2 7EF, UK

**Author notes:** Correspondence should be addressed to: Nitsan Schwarz, Sagol School of Neuroscience, Tel Aviv University, Tel Aviv 6997801, Israel.

## Abstract

**Background:** Age at menopause varies widely across women, yet little is known about how this relates to long-term behavioral and brain-structural changes. Previous research has focused primarily on the menopausal transition itself, and studies investigating cognitive outcomes suggest reduced age-related cognitive performance with earlier menopause.

**Objectives:** This study aims to investigate whether earlier age at menopause is associated with reduced cognitive performance and brain structure in later-life in a population-based cohort. To our knowledge, this is one of the only studies examining both cognition and neuroimaging in the same cohort of post-menopausal women, and the first study to formally test whether gray matter volume mediates the relationship between menopause timing and cognitive performance within the same population-based sample.

**Methods:** We analyzed data from the Cambridge Centre of Neuroscience and Aging, which included 747 postmenopausal women who underwent cognitive testing (Addenbrooke’s Cognitive Examine Revised, ACE-R). A subset (n=190) was additionally tested with a fluid intelligence test and underwent structural brain scans to measure gray and white matter volume (GMV and WMV). Multiple linear regression models were used to evaluate the association between menopause age and cognitive performance, as well as gray matter volume, controlling for chronological age.

**Results:** Earlier menopause was associated with lower cognitive performance, as measured by the ACE-R, with similar associations for fluid intelligence. Structural brain analyses revealed that earlier age at menopause was associated with decreased total gray matter volume (TIV-corrected). No significant interactions were observed between age at menopause and chronological age for any of the outcomes. GMV was a partial mediator between age at menopause and cognitive performance, while controlling for chronological age.

**Conclusion:** In a population-based sample, women with earlier age at menopause show both reduced cognitive performance and lower GMV, suggesting that GMV reduction may be one mechanism linking earlier menopause to cognitive decline. However, the cross-sectional nature of this study prevents causal conclusions, and longitudinal research is needed to establish causal links and to explore potential targeted interventions.

## Introduction

Menopause timing is increasingly recognized as an important factor influencing long-term brain health, but its effects on aging-related cognitive and structural brain changes remain poorly understood. With increasing life expectancy, understanding the role of menopause timing in cognitive and brain aging, as well as its contribution to sex-specific differences in brain health, is essential for targeted interventions.^1^

The menopausal transition is defined as the permanent cessation of ovulation and hence menstruation due to ovarian failure, and is influenced by numerous factors such as nutrition, exercise, number of pregnancies and childbirths, and socioeconomic background^2–4^. This transition ends in the postmenopausal stage, defined by 12 consecutive months without menstruation after which a woman’s ovaries do not release eggs and produce minimal estrogen^3,5^. In postmenopausal women, symptoms such as disturbed sleep, weight gain, and increased risks for osteoporosis and cardiovascular disease are common^6^. Cognitive health is also adversely affected, with studies linking menopause to age-related cognitive decline^3,7^. Moreover, recent studies suggest a relationship between menopause and accelerated brain aging, as evidenced by volumetric differences between postmenopausal and pre- or peri-menopausal women^8,9^.

Although general postmenopausal symptoms are widely known, less is understood about how the age at menopause affects these processes. Menopause generally occurs around age 51, with a typical range between 45 and 55 years^2,10^. However, approximately 10% of women experience early menopause (before age 45) and 1% of women experience premature ovarian insufficiency, where menopause occurs before the age of 40^11,12^. For women experiencing earlier menopause, particularly before the age of 45, the post-menopausal years come with increased risks compared to women with later menopause. These include a higher risk for cardiovascular disease, osteoporosis, and accelerated cognitive decline^5,6,13^.

Despite these findings, research on the relationship between menopausal timing and brain health remains limited in both quantity and quality. Existing research has explored the relationship between earlier menopause and cognitive performance, and earlier menopause and brain structure^14,15^. However, there are very few studies evaluating both within the same cohort, and no study to date has explicitly examined the mediation of this relationship by brain structure. This limits the ability to understand the mechanisms of earlier age at menopause and reduced cognitive performance later in life.

Our research aims to bridge this gap by examining how the timing of menopause relates to both long-term cognitive performance and brain structure. We hypothesized that an earlier age of menopause would be associated with both reduced performance in gross cognitive performance and reduced GMV within the same cohort of postmenopausal women, with GMV as the mediator between age at menopause and cognitive performance.

## Methods

### Participants

Data was obtained from the Cambridge Centre of Neuroscience and Aging, a large-scale research project launched in 2010 to investigate how individuals can best retain cognitive abilities into old age^16^. Participants were recruited from the general population within the Cambridge City area, UK, aiming to be representative of the UK population. All participants gave written informed consent prior to beginning the study. The study was approved by the Cambridgeshire 2 (now East of England - Cambridge Central) Research Ethics Committee, in accordance with the Declaration of Helsinki.

The project data was separated into stages, and the relevant stages for this study included: Stage 1, where 2591 adults aged 18-90 completed a home interview that included lifestyle questions and cognitive tests; and Stage 2, where approximately 700 participants underwent neuroimaging and additional cognitive testing^16^.

Out of the 2591 participants interviewed in Stage 1, 747 of the 1459 women reported being post-menopausal and were included in the analyses. Age at menopause was determined using the question, “How old were you when your periods stopped?”^16^, as done in previous studies.^17^

### Behavioral measures

During Stage 1, participants were interviewed in their home to acquire demographic information and measures of cognitive, mental and physical health. Cognitive assessment included Addenbrooke’s Cognitive Evaluation – Revised (ACE-R), which is a validated and widely used cognitive battery that assesses gross cognitive performance for dementia screening across five cognitive domains: Memory, Orientation/Attention, Fluency, Language, and Visuospatial ability^18^. Overall, 747 postmenopausal women completed the ACE-R.

A subset of participants from Stage 1 continued to Stage 2, based on additional exclusion criteria. Exclusion criteria for Stage 2 were described extensively in the original report^16^, including cognitive impairment (Mini-Mental State Examination scores below 25), difficulty in communication, substance abuse, mobility issues, significant medical issues, and MRI contraindications. In this stage, the relevant assessments included a fluid intelligence test and structural imaging. For fluid intelligence, The Cattell Culture Fair, Scale 2 Form A (Cattell) was used, which evaluates critical cognitive processes like fluid reasoning which decline with age^16,19,20^. The Cattell test is made up of four timed subtests with individual nonverbal “puzzles”, including Series Completion (3 minutes), Classification (4 minutes), Matrices (3 minutes), and Conditions (2.5 minutes). Participants mark the answers to each subtest using pen-and-paper in a multiple-choice format, for a maximum score of 46 (1 point for each correct question)^16^. A composite score was computed as the first principal component from the Principal Component Analysis (PCA) to the four subtest scores. ^19^. In total, 155 postmenopausal women had a complete Cattell dataset.

### Structural MRI

A 3T Siemens TIM Trio was used to assess white matter volume (WMV) and gray matter volume (GMV). Structural MRI data was acquired and preprocessed as described previously^20^. In short, T1-weighted image (MPRAGE, repetition time 2250 ms, echo time 2.99 ms, inversion time 900 ms, flip angle 9°, field-of-view 256 mm × 240 mm × 192 mm, isotropic 1 mm voxels) was acquired for each participant. Structural images were preprocessed using the automatic analysis batching system (http://imaging.mrc-cbu.cam.ac.uk/imaging/AA) in SPM12^20^. The images were segmented into gray and white matter using SPM12’s tissue priors, and we used total gray matter volume (GMV) and white matter volume (WMV) for the analyses. Total intracranial volume (TIV) was computed from the segmentation to account for differences in head size. In total, 188 postmenopausal women had valid structural MRI data.

### Statistical Analyses

To investigate the relationship between age at menopause and ACE-R score, a multiple linear regression model was used with age at menopause and chronological age, as well as their interaction as predictors. Each individual subdomain of the ACE-R was also examined in an exploratory analysis. Women who reported their last menstrual period at below 25 years of age (n=0) or above 60 years of age (n=9) were excluded due to a suspected incorrect response, as done previously^21^. Outliers in cognitive tests and GMV/WMV were defined as +/- 3 standard deviations from the mean and were excluded from analysis.

Three multiple linear regression models were conducted with Cattell test scores, GMV, and WMV as the dependent variables. GMV and WMV were adjusted to TIV to account for differences in head size. The independent variables were age at menopause, chronological age, and their interaction. MRIcron software was used for visualization of GMV and WMV (https://www.nitrc.org/projects/mricron).

A bootstrapped mediation analysis was conducted using multiple regression models to examine the effect of GMV on the relationship between age at menopause and cognitive performance. The mediation model was estimated with 5000 bootstrap resamples to obtain bias-corrected confidence intervals. All variables were Z-scored prior to the analysis. Chronological age was included as a covariate for all regression models.

All analyses were completed using Python version 3.10.0^22^.

## Results

We first evaluated our study sample in comparison to the global prevalence of age at menopause. The sample included 2.9% of women who would be classified as experiencing premature ovarian insufficiency and 7.9% who would be classified as early menopause, which is broadly consistent with global prevalence estimates^23^. Participant demographics can be seen in Table 1.

**Table 1.** Demographic data of Cam-CAN cohorts for each evaluation; ACE-R, Cattell and neuroimaging. Groups are split up into early and premature menopause (<45), normal menopause (45-55), and late menopause (>55) for organization purposes only. The rest of the data focuses on continuous age at menopause.

| Age at menopause ("Menopause") | ACE-R |  |  | Cattell |  |  | Structural Imaging |  |  |
| --- | --- | --- | --- | --- | --- | --- | --- | --- | --- |
|  | N | Menopausal Age Mean (SD) | Chronological Age Mean (SD) | N | Menopausal Age Mean (SD) | Chronological Age Mean (SD) | N | Menopausal Age Mean (SD) | Chronological Age Mean (SD) |
| <45 | 81 | 40.3(3.4) | 71.5(13.1) | 14 | 41.0 (2.7) | 64.9(12.6) | 20 | 40.3(3.1) | 69.8(13.4) |
| 45-55 | 600 | 50.7(2.9) | 73.6(11.6) | 127 | 50.8 (2.9) | 68.1(9.8) | 152 | 50.7(2.9) | 68.8(9.7) |
| >55 | 57 | 57.5(1.5) | 75.8(9.9) | 12 | 58.1 (1.4) | 76.6(7.5) | 16 | 58.2(1.5) | 74.6(9.3) |
| Total | 738 | 50.1(4.9) | 73.6(11.6) | 153 | 50.5 (4.6) | 68.4(10.2) | 188 | 50.3(4.9) | 69.4(10.2) |

### Study Sample

*Participant demographics are summarized in Table 1*.

### Cognitive Performance

We first conducted a multiple linear regression to test for the association between age at menopause, current age and their interaction on ACE-R score (Table 2). The model was significant at (*R*² = 0.22*, F*(2, 719) = 102.5, *p* < 0.001). There was no significant interaction between age at menopause and current age in predicting ACE-R (*t*(719) =-0.81*, p =* 0.42). Importantly, however, there was a significant association between age at menopause and ACE-R score (*t*(719) = 2.81*, p =* 0.005), indicating that earlier menopause was associated with lower cognitive performance, independently from current age. Specifically, a reduction of one year in the age at menopause corresponded to a 0.15-point decrease in ACE-R score (Fig. 1). The association remained significant, although weaker, when including education in the model.

**Table 2.** Results of the multiple linear regression analysis, examining the effect of age at menopause, current age, and their interaction on ACE-R score. Model 1 included the intercept, menopause age and age; model 2 included the interaction term Menopause Age*Age. All predictors are reported using Model 1, except for the interaction term (Model 2).

| Predictor | Estimate | SE | t-statistic | p-value |
| --- | --- | --- | --- | --- |
| Intercept | 108.31 | 3.92 | 27.61 | <0.001 |
| Menopause Age | 0.19 | 0.07 | 2.81 | 0.005 |
| Age | -0.39 | 0.03 | -14.20 | <0.001 |
| Menopause Age*Age | 0.006 | 0.004 | -1.60 | 0.11 |

**Figure 1.**
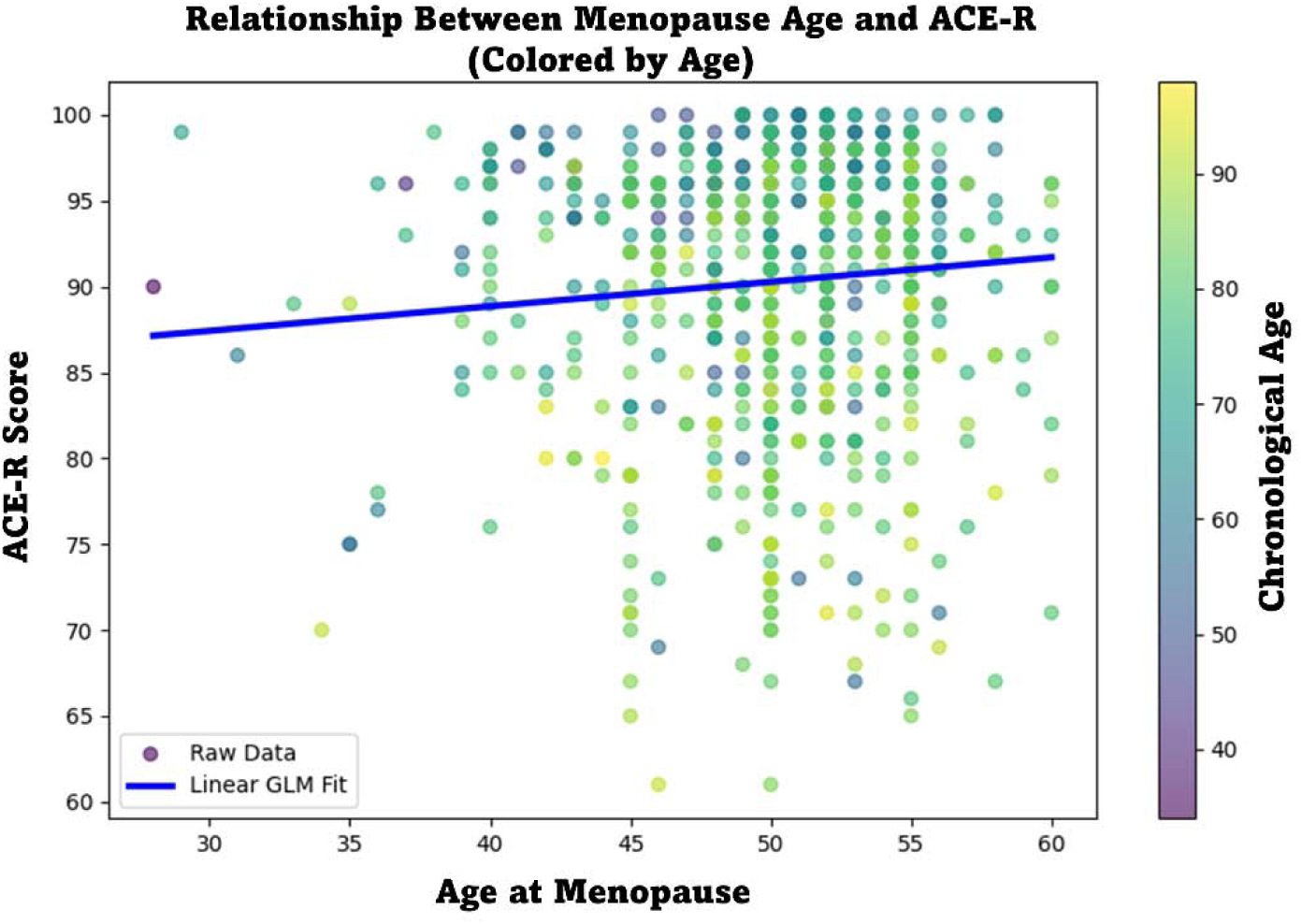
Generalized linear model demonstrating relationship between menopause age and ACE-R score, with chronological age shown.

Exploratory, domain-specific analyses showed that Language and Visuospatial domains followed the same trend as overall ACE-R score (Supplementary Material). Similar results were also found for the Cattell (Supplementary Material).

### Structural imaging

After showing an association between two independent cognitive tests and age at menopause, we investigated the relationship between brain structure and age at menopause. We conducted a linear regression analysis on GMV and WMV, adjusted to total intracranial volume, to see if this trend could also be seen in brain structure. The model for GMV was significant (*R*^2^=0.22, *F*(2, 182)= 24.53, *p* < 0.001). To account for individual differences in head size, we analyzed GMV as the ratio between GMV and total intracranial volume (TIV), as typically done (Table 3)^24^. The results suggest that an earlier age at menopause was significantly associated with lower GMV independently of age. Specifically, a reduction of one year in the age at menopause corresponded to a 0.0014 decrease in the GMV to TIV ratio, or 2370mm^3^ in total GMV (Fig. 2). The model with white matter volume as the dependent variable showed no significant association with age at menopause (*t*(182) =-0.20*, p =* 0.84)

**Figure 2.**
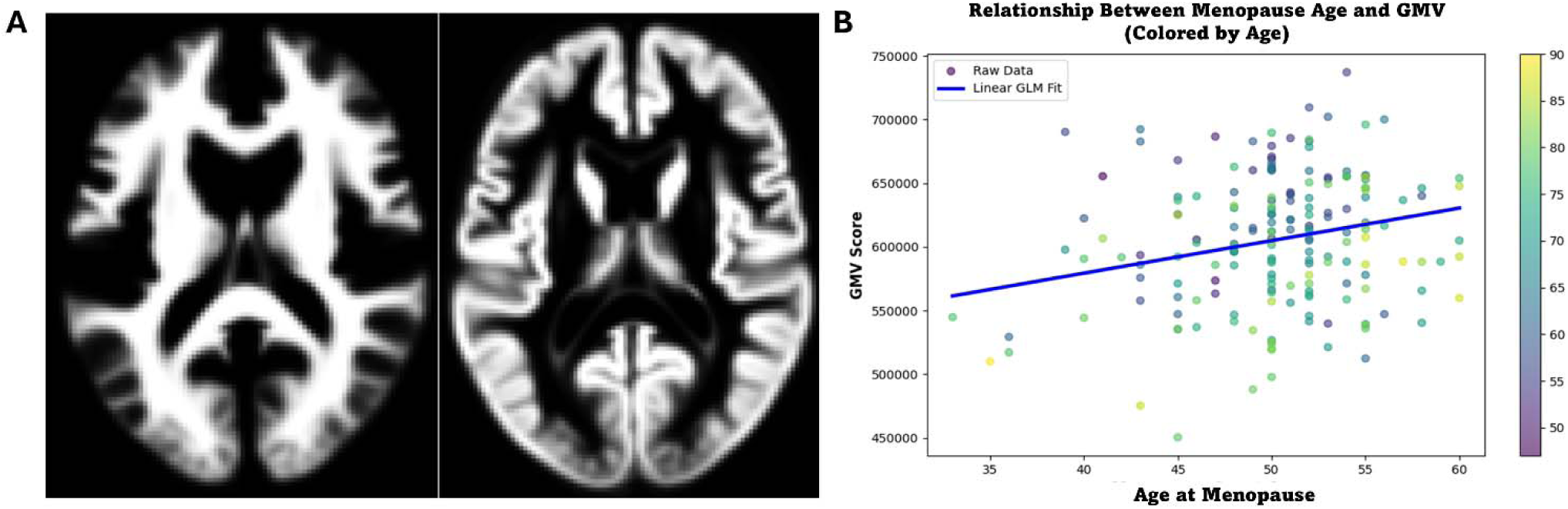
A) Horizontal sections of white matter (left) and gray matter (right), as computed during segmentation. B) Generalized linear model demonstrating relationship between menopause age and gray matter volume (GMV), colored by chronological age.

**Table 3.** Results of the multiple linear regression analysis, examining the effect of age at menopause, chronological age, and their interaction on GMV (as GMV to TIV ratio to account for TIV). Model 1 included the intercept, menopause age and chronological age; model 2 included the interaction term Menopause Age*Age. All predictors are reported using Model 1, except for the interaction term (Model 2).

| Predictor | Estimate | SE | t-statistic | p-statistic |
| --- | --- | --- | --- | --- |
| Intercept | 0.51 | 0.0222 | 23.11 | <0.001 |
| Age at menopause | 0.001 | 0.000385 | 2.86 | 0.005 |
| Age | -0.002 | 0.000186 | -8.24 | <0.001 |
| Interaction | $8.08 \times 10^{-5}$ | $3.4 \times 10^{-5}$ | 2.23 | 0.03 |

**Figure 3.**
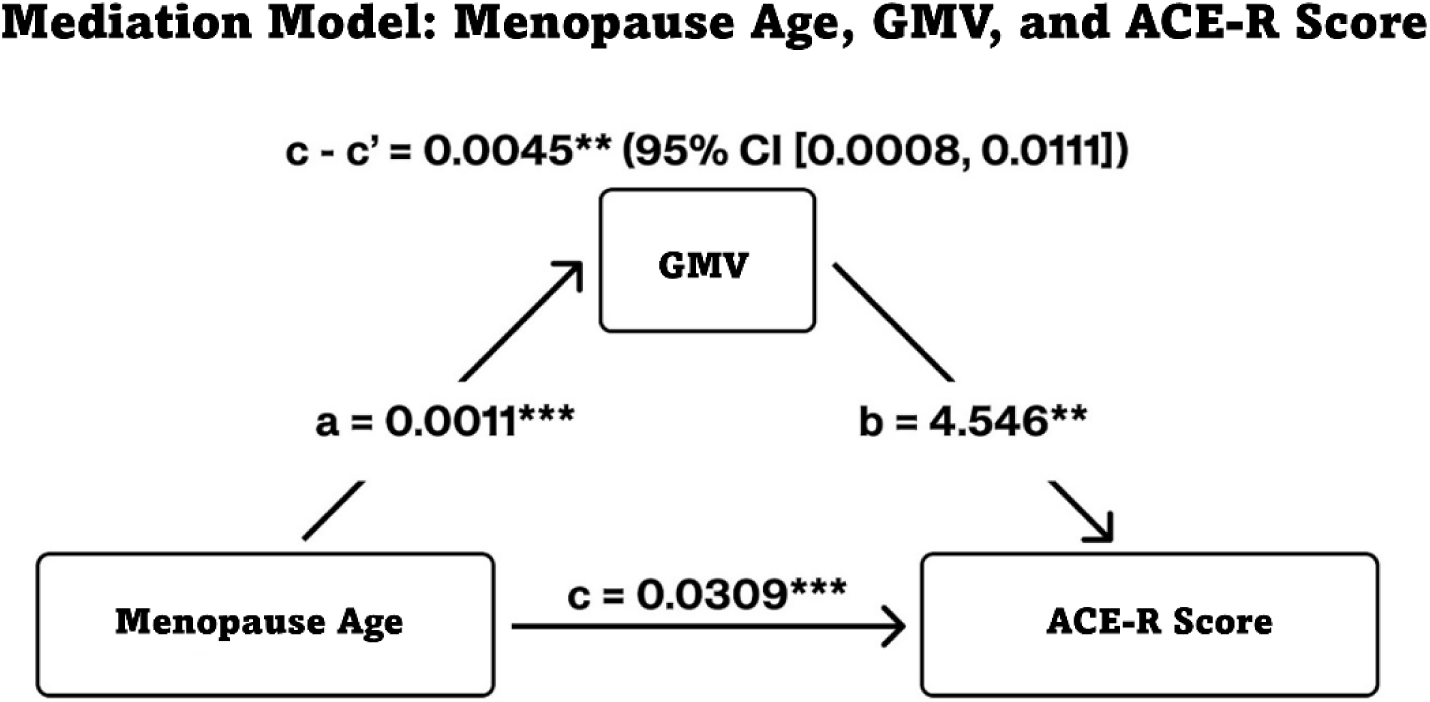
Mediation model of menopause age, GMV, and cognitive performance (ACE-R Score). A mediation model illustrating the relationships between menopause age, GMV, and ACE-R Score. The numbers next to the arrows represent standardized path coefficients. Solid arrows indicate statistically significant paths (*p < 0.05, **p < 0.01, ***p < 0.001). The indirect effect (c - c’) and its confidence interval are reported at the top.

We conducted an exploratory mediation analysis to examine whether GMV statistically mediates the relationship between menopause age and cognitive performance (ACE-R score), while accounting for age. In the GLM, age at menopause significantly predicted GMV (path *a* = 0.001, *p* = 0.004), and GMV significantly predicted ACE-R (path *b* = 4.546, *p* = 0.003). The total effect of menopause age on ACE-R was significant (*c* = 0.031, *p* < 0.001). The indirect effect (*c* - *c*’ = 0.005, 95% CI [0.0008, 0.011]) was significant (*p* = 0.006). The direct effect remained significant after accounting for GMV (*c*’ = 0.026, *p* < 0.001), suggesting that GMV explains part of the association between menopause age and cognitive performance.

## Discussion

While the few studies in the field have looked at cognition and neuroimaging separately, our study is unique in that it evaluates both cognition and brain structure in the same population-based, cross-sectional cohort of postmenopausal women. Our findings demonstrate a significant relationship between age at menopause and both reduced cognition and GMV, independent of current age. The mediation analysis, which is more limited in a cross-sectional context, suggests that women with earlier menopause show both reduced cognitive performance and gray matter volume.

Both the ACE-R and fluid intelligent assessments showed a significant negative association with earlier age at menopause. These results are consistent with previous studies showing associations between cognitive decline and women experiencing premature, early and earlier menopause^3,25,26^. Moreover, our findings add to this line of research by linking earlier menopause to fluid intelligence performance, which is highly sensitive to aging^19^. These findings suggest that women who experience earlier menopause may exhibit signs of accelerated cognitive aging, with lower cognitive performance later in life. However, the lack of a significant interaction between menopause age and chronological age indicates that while earlier menopause is linked to lower cognitive function, it does not appear to alter the rate of age-related cognitive decline. Longitudinal research in the field can specifically test these hypotheses.

Interestingly, exploratory analyses on the subdomains of the ACE-R suggested that a specific reduction in Language, Visuospatial and Orientation/Attention domains were most strongly associated with age at menopause. Research has suggested that language and visuospatial abilities in old age are specifically predictive of vascular dementia risk ^27^. Our findings that earlier menopause is associated with a reduction in the scores of the ACE-R subdomains that are associated with vascular dementia (Language, Visuospatial) reflect an urgent need for further sex-specific research in dementia mechanisms; especially as estrogen exposure, which is known to drop dramatically during and post-menopause, has been linked specifically to vascular dementia risk^28^.

In terms of differences in brain structure, earlier menopause was significantly associated with reduced GMV, independently of age, suggesting that menopause timing has a measurable and lasting impact on brain structure. Differences in GMV statistically mediated the relationship between age at menopause and cognitive performance, indicating that GMV may play a role in linking early menopause to cognitive decline. Future studies should investigate the temporal dynamics between menopause timing, brain structure changes, and cognitive decline, as well as explore potential interventions to mitigate these effects. While lower GMV may be one mechanism linking early menopause to reduced cognitive performance, our mediation model provides the first statistical support for this hypothesis. Nevertheless, this finding requires confirmation through longitudinal research.

The magnitude of GMV loss, in a clinical perspective, holds implications for aging: Across an 8-year difference in menopause timing (e.g., menopause at 44 vs. 52), this amounts to an estimated 2.6–2.7% reduction in total gray matter volume, based on an average total GMV of 740,000 mm³ in healthy adult women^29^. This corresponds to approximately 5–8 years of normative brain aging, based on normative estimates of GMV loss in typical ageing^30^.

GMV loss is known to be associated with increased risk of dementia, which suggests that earlier menopause could predispose women to structural brain changes that contribute to dementia risk later in life and possibly vascular dementia in particular^31^. Notably, there was no association between menopause age and white matter volume, further suggesting that menopause influence on brain structure is specific to changes in GMV. However, future research should also use complementary techniques, such as diffusion weighted imaging.

Mechanistically, these findings align with both human and animal research theories that estrogen exerts neuroprotective effects^28^. Estradiol, particularly, does this through modulation of synaptic plasticity and neuronal health, and its abrupt decline during menopause is hypothesized to play a key role in accelerated brain aging^3,4,14^. Recent theories propose that earlier menopause could lead to a longer duration of low estrogen exposure after midlife, possibly contributing to decline of cognition and GMV^28^. While it is important to note that our study did not measure levels of ovarian hormones in our cohort, the knowledge of estrogen decline post-menopause and sex-specific disproportion in dementia demands more research into how ovarian hormones play a part in dementia prediction. Hormone replacement therapy (HRT), specifically, is known to reduce the risk of cognitive decline when initiated during the menopausal transition, and potentially mitigate the negative effects of estrogen depletion^3,28^. However, the timing of HRT is critical, as seen by the “critical window” hypothesis which suggests that HRT is neuroprotective only when administered within five years of menopause^32,33^, and that administration of HRT in older post-menopausal women could even be detrimental to cognitive health^34,35^.

### Strengths and Limitations

A key strength of this study is the integration of cognitive and fluid intelligence assessments, as well as neuroimaging, within the same cohort of postmenopausal women. Existing research focuses separately on either cognitive assessments or neuroimaging, and so this integration allows us to draw connections between these critical domains. By combining both approaches, our study describes a more comprehensive understanding of how menopause timing influences long-term brain health. Given the inconclusive findings on early menopause, cognition, and neuroimaging, our results urge a greater look at menopause timing studies relating to cognition and neuroimaging, and as a mediator of GMV.

In terms of limitations, we did not collect data on HRT use. The lack of HRT data prevents us from distinguishing between natural age-related changes and those potentially attenuated by hormonal intervention. Future research should explicitly account for HRT use to understand its timing effects on both cognition and GMV. Another limitation is the exclusion of participants with low cognitive performance from the fluid intelligence and neuroimaging analyses. This exclusion criterion may have inadvertently biased our sample by removing those most vulnerable to cognitive decline, therefore limiting the generalizability of our findings to the broader population of postmenopausal women. Additionally, while the statistical mediation model suggests GMV may link earlier menopause and cognitive performance, the cross-sectional design prevents any inference of causality or temporal ordering.

## Conclusions

Our findings reveal significant associations between earlier menopause and both reduced cognitive performance and GMV, independently of age, in the same cohort of postmenopausal women. The results suggest that GMV reduction may be one mechanism linking earlier menopause to cognitive decline. Longitudinal studies are highly needed to establish causal links and to explore potential targeted interventions to mitigate the possible detrimental effects of earlier menopause.

## Data Availability

All data produced in the present study are available upon reasonable request to the authors.

## Supplementary Materials

### ACE-R Subdomains

Domain-specific analyses showed that Language and Visuospatial domains, linked to vascular dementia in literature ^27^, followed the same trend as overall ACE_R score, with Language ( *t*(719)= 2.64, *p*=0.005), Visuospatial (*t* (719)= 2.06, *p*=0.040), and Orientation/Attention (*t*(719)= 2.14, *p*=0.033) domains significantly associated with age at menopause. The other domains (Fluency and Memory) showed no significant relationship with age at menopause (Table 4). This is visualized in Fig. 4.

**Table 4:** Multiple linear regression association between ACE-R domains and age at menopause (“Menopause”) with chronological age as a covariate. Results significant to age at menopause are highlighted.

| Subdomain | Predictor | Estimate | SE | t-statistic | p-statistic |
| --- | --- | --- | --- | --- | --- |
| Visuospatial | Intercept | 1.05 | 0.0459 | 22.81 | <0.001 |
| | Menopause | 0.002 | $8.17 \times 10^{-4}$ | 2.06 | 0.04 |
| | Age | -0.003 | $3.41 \times 10^{-4}$ | -8.18 | <0.001 |
| Language | Intercept | 1.00 | 0.0339 | 29.54 | <0.001 |
| | Menopause | 0.002 | $6.04 \times 10^{-4}$ | 2.64 | 0.008 |
| | Age | -0.002 | $2.52 \times 10^{-4}$ | -7.30 | <0.001 |
| Fluency | Intercept | 1.20 | 0.0676 | 17.74 | <0.001 |
|  | Menopause | <0.001 | 0.00120 | 0.49 | 0.62 |
| | Age | -0.006 | $2.52 \times 10^{-4}$ | -10.89 | -0.006 |
| Memory | Intercept | 1.15 | 0.0553 | 20.73 | <0.001 |
| | Menopause | 0.002 | $9.84 \times 10^{-4}$ | 1.63 | 0.12 |
| | Age | -0.005 | $4.11 \times 10^{-4}$ | -11.81 | <0.001 |
| Orientation/Attention | Intercept | 1.00 | 0.0358 | 27.86 | <0.001 |
| | Menopause | 0.001 | $6.37 \times 10^{-4}$ | 2.14 | 0.03 |
| | Age | -0.002 | $6.22 \times 10^{-4}$ | -6.50 | <0.001 |

**Figure 4:**
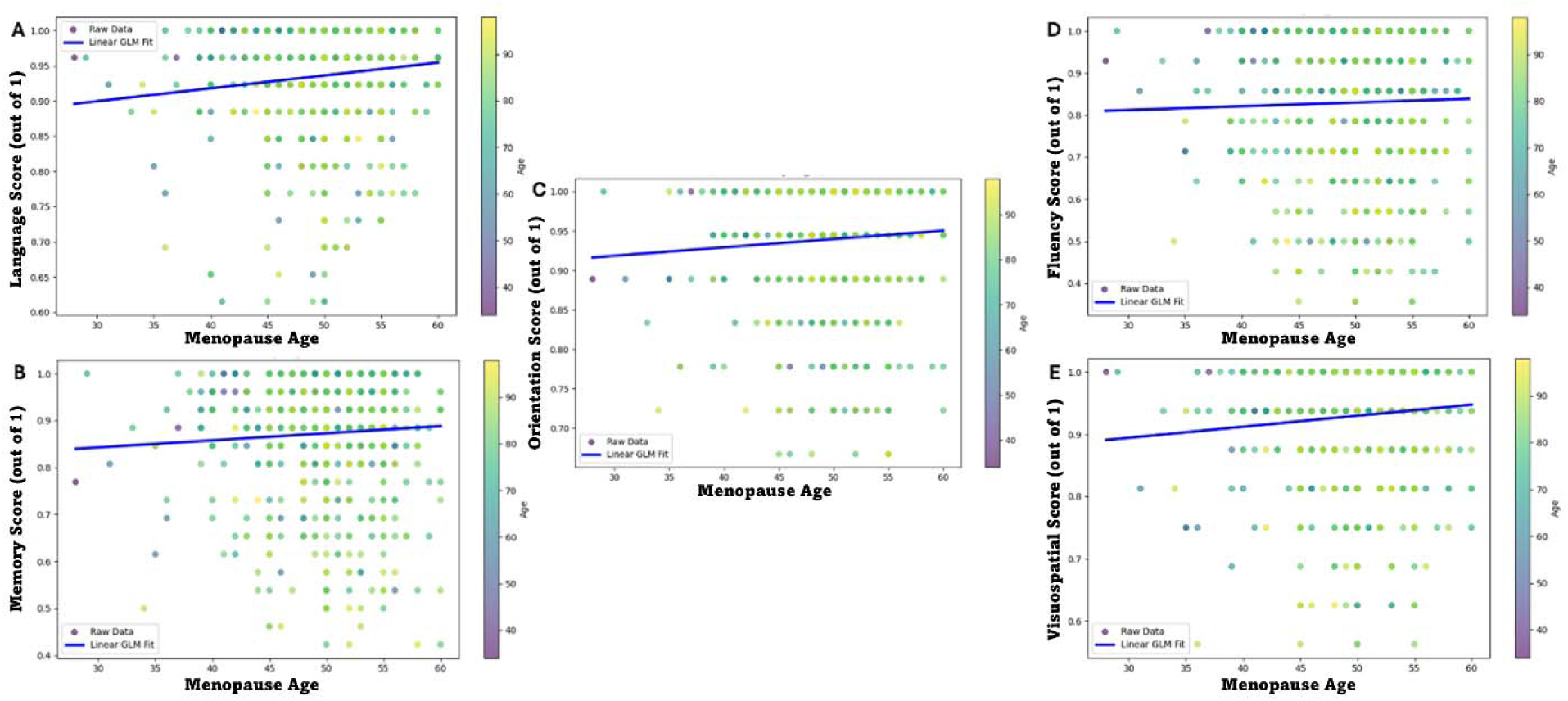
*GLM Plots for ACE-R Subdomains.* The plots illustrate generalized linear model relationships between age at menopause and performance in five cognitive domains of the ACE-R: Language (A), Memory (B), Orientation/Attention (C), Fluency (D) and Visuospatial (E) with chronological age visualized by color.

### Fluid Intelligence: Cattell Test

In a subset of participants who completed Stage 2, we investigated whether a relationship with age at menopause would be found for performance in the Cattell test, such as that found for ACE-R. The model was significant (*R*^2^= 0.35, *F(2, 146)*= 38.45, *p*<0.001), and there was a small but significant association between age at menopause and Cattell score, above and beyond current age (*t*(146) = 3.116, *p*=0.002) (Table 3). Specifically, a reduction of one year in the age of menopause corresponded to a 0.15-point decrease in the Cattell score (Fig. 5). A robust regression evaluation remained significant (*p*= 0.010).

**Table 5.** Results of the multiple linear regression analysis, examining the effect of age at menopause, current age, and their interaction on Cattell score. Model 1 included the intercept, menopause ageand age; model 2 included the interaction term Menopause Age*Age. All predictors are reported using Model 1, except for the interaction term (Model 2).

| Predictor | Estimate | SE | t-statistic | p-statistic |
| --- | --- | --- | --- | --- |
| Intercept | 19.49 | 2.51 | 7.77 | <0.001 |
| Menopause Age | 0.15 | 0.0470 | 3.12 | 0.002 |
| Age | -0.18 | 0.0210 | -8.66 | <0.001 |
| Interaction | 0.004 | 0.00430 | 0.92 | 0.36 |

**Figure 5:**
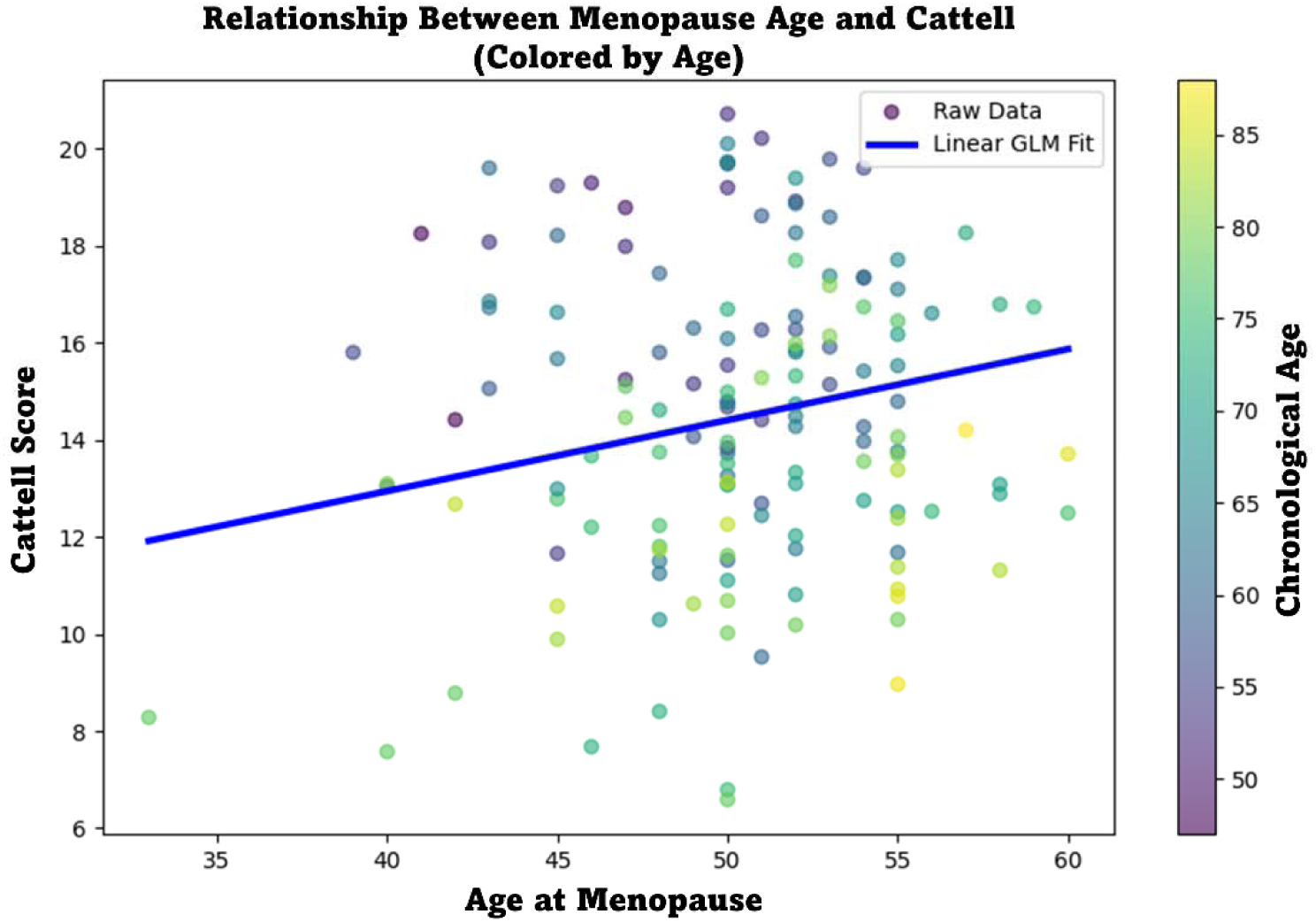
Generalized linear model demonstrating relationship between age at menopause and ACE-R score, with chronological age shown.

